# Implementation of Indoor residual spraying for malaria vector control in Uganda: An exploration of community experiences

**DOI:** 10.64898/2026.01.15.26344175

**Authors:** Moses Ocan, Nakalembe Loyce, Josephine Bayiga, Geofrey Kinalwa, Emmanuel Arinaitwe, Henry Mawejje, Rogers Naturinda, Moses Adriko, Simon Kigozi, Sam Nsobya

## Abstract

**Background:** Malaria vector control using indoor residual spraying (IRS) is a key intervention in the fight against malaria globally. However, IRS is faced with several challenges limiting wide scale deployment especially in sub-Saharan Africa. This study explored community experiences on implementation of IRS using clothianidin-deltamethrin, and pirimiphos-methyl insecticides in west Nile region of Uganda.

**Methods:** Using a phenomenological design were individuals involved in the implementation of IRS were purposively included in the study. In-depth interviews were conducted using an interview guide. The interviews were conducted in both the local language of the area (Aringa) and English whenever appropriate. All interviews were audio recorded and transcribed verbatim. The transcripts were then analyzed using thematic analysis using Nvivo *ver 14.0* software.

**Results:** A total of 30 in-depth interviews were conducted. Majority, 60% (18/30) of the study participants were females. The themes that emerged included coverage of the household with the spray, practices of stakeholders, perceptions regarding insecticides, opportunities, and challenges facing implementation of IRS. Perception on effectiveness, involvement of different stakeholders including local leaders, religious leaders and training of spray teams helped enhance acceptance. While smearing or covering of sprayed walls, lack of transparency in recruitment and payment of spray operators, removal of house items, and shortage of spray supplies threaten scaling up of IRS implementation.

**Conclusion:** Communities were highly receptive towards IRS for malaria control with most study participants being positive on its effectiveness. Training of spray operators, involvement of different stakeholders, community compliance with local authorities, and involvement of social behavior change communication (SBCC) help enhance implementation of IRS. However, the deployment faced some challenges including inadequate supplies, removal of house items, and difficulty moving spray materials in the field. The Ministry of health and implementing partners should prioritize strengthening timely community engagement prior to IRS deployment.

## Introduction

Indoor residual spraying (IRS) remains a key malaria vector control measure and is responsible for the gains in malaria elimination efforts globally (1, 2). In most malaria endemic countries, especially sub-Saharan Africa, IRS is implemented alongside mass bed net distribution and chemoprevention for malaria control (3, 4). Additionally, in Uganda bed nets are routinely distributed to pregnant women during antenatal and immunization clinic visits (5). However, despite implementation of different interventions, malaria burden remains high in sub-Sahara Africa accounting for over 95% of malaria cases worldwide (2). For example, in Uganda malaria is still responsible for 30-50% of outpatient health facility visits, 35% of hospital admissions and 9-14% of hospital deaths especially among children under five years (6). This indicates potential stalling of malaria control measures including IRS and could be due to several factors including malaria vector insecticide resistance, community uptake, and change in vector behavior (4, 7).

The implementation of IRS faces several challenges both at household and national level in most malaria affected settings. The complex logistics associated with IRS deployment limits wider application with most countries only deploying it in high malaria transmission settings (2, 4). In addition, the frequency of application is also affected with most countries only managing one cycle of IRS deployment annually irrespective of residual efficacy of the insecticide (8). In 2014, Uganda scaled up its IRS deployment to 14 districts including both epidemic prone (southwest) and high malaria burden districts (eastern and northern) (9). Indoor residual spraying is irregularly deployed in Uganda for example between 1960 to early 2000 its application was ceased and only relaunched in 2006 (8). The intermittent application potentially affects the overall contribution of IRS to malaria control in the country.

The change in malaria mosquito vector population and behavior has also been reported in most settings in sub-Saharan Africa(10–12). The current IRS deployment targets indoor resting mosquitos, however, studies have reported increased outdoor biting among mosquito vectors(7, 8) (Mwesige et al., 2025; WHO malaria report, 2006). This coupled with delays in entering the house in the evening where individuals stay outdoors late into the night further increases the risk of mosquito bites. A recent study reported *Anopheles gambiae* and *Anopheles funestus* as the two most prevalent malaria vectors in Uganda(7). While the insecticides used in IRS remain efficacious, studies are reporting widespread emergence of resistance among mosquito vectors(13). This coupled with increased outdoor biting and delay in entering the houses in the night could potentially be contributing to decreased effectiveness of IRS in malaria elimination in sub-Saharan Africa(14).

The extent of uptake of indoor residual spraying for malaria control varies in different communities across sub-Saharan Africa(2). While IRS deployment remains largely accepted across most malaria endemic settings, studies have reported instances of reduced uptake of the spray by household members(14). This could potentially be due to inadequate community engagement coupled with the complex IRS implementation logistics(4). The optimal community-level protective effect of IRS has been reported to occur at a coverage greater than 80% as recommended by the World Health Organization (15). From previous studies individual and household protection from IRS has been reported to be derived from community level coverage(16, 17). Studies in most sub-Sahara African countries have found lower community level IRS coverage (17). Recent IRS campaigns in Uganda have reported coverage of over 80% however, identifying the houses that should be included in the spray coverage denominator presents a challenge and thus the true IRS community coverage remains unclear.

West Nile region first received IRS in November/December 2022 with Clothianidin-deltamethrin and in December/2023 with Pirimiphos-methyl and subsequently with pirimiphos-methyl. Despite repeated application of IRS and implementation of other control interventions, Uganda remains among the top three high malaria burden countries contributing 5% of the global malaria cases (2). The persistence of malaria in the country could be an indicator of the challenges in the implementation of different malaria control interventions. This study explored experiences of communities on the implementation of IRS in Yumbe district, West Nile region a high malaria transmission setting in Uganda and thus help guide policy and decision makers in designing context specific strategies to help improve acceptance and increase community IRS coverage.

## Materials and methods

### Study design and settings

This was a phenomenology study conducted in Yumbe district a high malaria transmission setting in West Nile region of Uganda. Since 2022 the district has had five cycles of indoor residual spraying with two different chemicals (first IRS cycle used Clothianidin-Deltamethrin and was deployed in November/December 2022, and Pirimiphos-methyl was used in the second and subsequent IRS cycles starting on December 2023). The national malaria elimination division of the Ministry of health implemented the indoor residual spraying of the entire district in partnership with the President Malaria Initiative (PMI). Data collection was done between February and April 2024. Packaging of the IRS insecticides and the brand names were used by communities in describing the different IRS cycles. For clothianidin-deltamethrin it was described using the sachet or Fludora Fusion while bottle or Actellic for pirimiphos-methyl.

### Study population and sampling

Data collection was done among community leaders, IRS field supervisors, IRS spray operators, IRS team leaders and household heads. Individuals who were involved in the deployment of IRS in communities were purposively selected for inclusion into the study.

### Data collection procedure

Data was collected using in-depth interviews following an interview guide. The In-depth Interview guide was developed using information from previous studies (14, 18). The tool was reviewed by members of the integrated vector management in the National malaria control division of the Ministry of Health. Information from the review was used to adjust the final interview guide. The IDI interview guide collected information on the following areas i) socio-demographic characteristics, ii) knowledge on indoor residual spraying, iii) facilitators, and iv) barriers/challenges of indoor residual spraying for malaria control. The interviews were conducted by two reaserch assistants (OW and IT) both of whom had bachelors degree training in social science and experienced in qualitative data collection. The interviews were conducted in the local language (Aringa) and english whenever appropriate and lasted on average 60 minutes. The research assistants were trained on the study proposal and ethical conduct of research prior to field data collection. A written informed consent was obtained from each participant prior to data collection. The interviews were audio recorded using SONY recorder (SONY Corp, China) and additional field notes collected during each of the interviews. The research assistants probed for further details relevant to each interview question inorder to gain more insight into the area of discussion. The discussion areas (topics) further evolved through the process of the fieldwork, both within each interview and from one to the next. Summaries of the field notes were written after each day of data collection and discussed within the field data collection team, to identify any new areas emerging for exploration in subsequent interviews. The interviews were conducted until information saturation was reached.

### Data management

After each data collection day, the audio files were downloaded onto a password protected computer hard drive and backed up. The audio recordings were listened to carefully and then transcribed verbatim into Word in english. For those in the local language, the transcripts were then back translated into Aringa to ensure that the meaning is maintained. The transcripts were then exported to NVivo *ver 14.0* (QSR International, Cambridge, MA) software for coding and analysis (19). A standardised format was applied to all transcripts to facilitate the comparison of data at the analysis stage. This included a summary of quantitative data that described participant’s demographic characteristics, the location and other key information to situate the interview. For this study, the transcription method reflected the interpretative approach. This included word-for-word transcription, recording all hesitations, pauses, utterances, cross-talking and incomplete sentences. Major interruptions by other people or telephones were recorded to contextualise any breaks in speech or repetitions. The transcription was proof-read against the audio file by the lead researcher (OM) to check for accuracy. Translation from aringa (local language) to english took a meaning-based approach from the original language into English. The translator (IT) was familiar with the research and its objectives. Quality criteria for the translations was comprehensibility (especially relating to culture specific concepts), appropriateness (in content and approach) and accuracy (faithful to the source text and key facts). Back-up files of word and NVivo documents were stored daily on an external encrypted hard drive.

### Data analysis

Interview transcrpts were analysed using a coding scheme developed from the themes emerging from the data. Prior to the coding process, a coding tree was developed listing all the identified themes and sub themes. The transcripts was reviewed and all statements pertinent to the sub themes were identified and were extracted. A total of f 30 transcripts were analyzed inductively, and NVivo *ver* 11.0 software was used to organize and interpret the data. The data were coded by two qualitative analysts (JB and AT). The initial coding was conducted independently by the lead social scientist (AT), discussed with the lead researcher (OM), a final coding scheme was then applied to all transcripts. Alongside this coding, a reflective analytical diary were kept, to draw out and justify any emerging themes. The analysis focused on facilitators, barriers and participant knowledge of IRS in the communities. Statements (quotes) from respondents are included for illustration to help contextualise the study findings.

### Ethical considerations

The study proposal was reviewed and approved by the Makerere University School of Biomedical Science Research Ethics Committee (SBS-2023-456). Research clearance to conduct the study in Uganda was obtained from Uganda National Council of Science and Technology (HS341ES). Administrative clearance was also be obtained from study districts and hospitals. A written informed consent was obtained from study participants prior to the interviews.

## Results

### Characteristics of study participants

A total of 30 respondents were included in the study. Of these, majority 60% (18/30) were females. The average age of the study participants was 36 years. The participants included spray operators (9), field team leaders (7), household heads (7), field supervisors (5), community leaders (2).

### Themes that emerged following analysis of in-depth interview data

From the interview data analysis, five themes emerged and these include.

1. Coverage of the household with the spray
2. Practices of stakeholders in the implementation of indoor residual spraying
3. Opportunities facing implementation of indoor residual spraying
4. Challenges facing implementation of indoor residual spraying
5. Perceptions regarding the insecticides used for indoor residual spraying

### Coverage of households with IRS for malaria control

In each household the spray operators focused only on houses were people sleep. The study participants highlighted that food storerooms, kitchens, bathrooms, and any other places where people do not sleep like latrines, and the compound, were not sprayed. In indoor residual spraying, the insecticide is deposited on the interior surfaces of walls of residential houses to kill indoor resting malaria mosquitos. The deployment of insecticides reported by respondents in this study is consistent with WHO recommendations(2). However, recent reports on outdoor biting of *An. funestus* and *An. gambiae s.s.* in IRS settings could potentially affect the effectiveness of IRS in malaria control (7).

> *“In my home all the houses were sprayed, but the latrine was not and you know mosquitoes like dark places. So, they did not reach our latrines, but the rooms where people sleep were all sprayed*.” --IDI-02-Household Head.
>
> *“Sitting rooms…yeah, we sprayed them…because you know people they stay long, they take long in sitting rooms. So, we spray sitting rooms and then bedrooms but the stores, we don’t spray them, where somebody cannot sleep, we can’t spray…”* --IDI-15-Team Leader of Spray Operators
>
> *“They[sprayers] sprayed my house the only thing that they did not spray it’s the kitchen and the toilet…they[sprayers] educated us seriously that the houses that are needed to be sprayed are those for sleeping, the one that are not supposed to be sprayed are kitchen and the one that food stuffs are being kept into it, the third one its latrines that are not supposed to be sprayed…the outside wall they did not spray”* -- IDI-27-Household Head.

Complaints were raised by some households about the presence of mosquitoes after spraying of the IRS chemicals which caused doubt about quality of the spray. Following complaints from some individuals in the community, the spray operators returned to some of the houses with complaints or those which were not unsprayed and re-applied the spray.

> *“But there are some complaints…some people, they told us they spray under dose, that was the only challenge there. But the rest of the people are saying the medicine is okay. But some few people in the village, they are complaining with the Sachet medicine (Fludora Fusion) …if the people don’t mix the medicine properly…then the chemical will remain in the tank…”* -- IDI-05-Spray Operator.
>
> *“…you make sure you go back to spray…, you ask the household, was the mixing done well? that is now the information we get from households…they also help us to monitor the execercise…”* IDI-08-Team Leader of Spray Operators
>
> *“After the spraying exercise was done we were also able to move to those villages where the spraying was done and find out people’s concerns and most of the people were able to give us positive responses about the intervention. So, they were able to request for more intervention to be in the place*”-- IDI-22-Supervisor.

### Practices of stakeholders in the implementation of indoor residual spraying in West Nile region, Uganda

#### Mixing of chemicals for indoor residual spraying

Appropriate mixing of the insecticides in the spray tanks is key to help ensure adequate concentration of the insecticide prior to spraying of the walls. Nearly all spray operators interviewed in this study were conversant with mixing procedures for the IRS insecticides that were used. They mentioned that for the insecticides in the sachets that were used in the 2022 cycle, start by pouring it in about 3 liters of water, mix, and then topped up to make a maximum level of 7.5 liters per sachet in the spray tank. Only two respondents stated that one sachet was mixed with up to 8 liters of water.

> “*Mixing of the chemicals are, you when you are mixing that chemical in the Sachets (Fludora Fusion) first you pour, 3-litre water in the pump, after which you remove the sachet inside there, you either dip it in that pump and you close it, after which you shake the pump. You shake the pump, aah after which it dissolves. You shake it vigorously, 5 times… It dissolves there, after which you open and you also add another, water which equivalent to 7.5 litres. After which you also shake it again before you go inside the house to spray*” --IDI-21-Team Leader of Spray Operators

Some of the respondents reported that the process of mixing the insecticide that was packed in Sachets (Fludora Fusion) was difficult, leading to sprayers not mixing it properly. Improper shaking caused the chemical to settle in the tank.

> *“They [people in the community] said that our chemicals were not good… some sprayers used to spray without shaking properly, the chemical remains down…, then while they are spraying it just remains in the tank. Because they told us when you are spraying, you must shake… you shake the pump… the chemical will not settle down… some spray operators didn’t shake…”* --IDI-17-Team Leader of Spray Operators.

However, for the insecticide packaged in bottles (Actellic) that was used in 2023 and subsequent cycles of IRS, most participants stated that it was easy to mix compared to the one in sachets (Fludora Fusion). The IDI participants reported preference for bottle IRS chemical over Sachets due to the ease of mixing.

> *“I was saying they should continue to use this recent one… because that one is just in the bottle (Actellic)..you can just pour it in the spray tank. But the one in the sachet (Fludora Fusion)…like you forget to shake very well … can not easily mix… But for this one in the bottle, you just pour…”* --IDI-10-Spray Operator.

#### Protection of individuals in households during spraying of IRS insecticides

The insecticides used for indoor residual spraying have potential to cause adverse effects to individuals who get in contact(18). During the spray exercise, the spray teams reported providing instructions to individuals in households to help protect them from potential exposure to the insecticides. Members of the households were requested to stay outside of the house during the spray exercise. Individuals were also instructed by the spray team to bring out all house items and cover them including water points with polythene bags. The polythene bags for covering house items were provided by the spray teams. This was done to help protect individuals from getting exposed to the insecticides. Members of the household also helped with provision of water to the spray teams for mixing the IRS insecticides.

> *“The household member is the one who brought for the water because you tell that person that I’m coming to mix this drug here and then you can bring for me like 10 liters of water. You mix the chemical (insecticide) when the people are seeing”* -- DI-03-Team Leader of Spray Operators.
>
> *“…Because what we do was to advise the household during the time of spraying…bring items out of the house and cover all including food stuffs and water points using something like the carpet or polythene bag…”* -- IDI-13-Spray Operator.

The study participants reported that during the spray exercise, children were not allowed to enter the house while the spraying was ongoing which helped protect them from getting exposed to the insecticides.

> *“The children we decided to keep them in the houses that were not supposed to be sprayed, and we collected to them food so that they can eat from there, we closed the doors and I can’t remember if how many hours we opened it later and it did not affect the children”* -- IDI-27-Household Head.

Household occupants were asked not to lean on the wall after spraying, to close the door for some time before opening, clean the household, dispose the residuals in a pit latrine, and not to hang items on the wall.

> “*After spraying, I went out, have told him also let a house close the door, let the house stay 2 hours minus opening after 2 hours, you open the windows, doors then aeration will be 30 minutes for aeration…”* --IDI-06-Team Leader of Spray Operators.

#### Documentation, and marking of sprayed houses

Documentation is key as it helps in monitoring implementation of IRS in communities. All information was captured by the spray operators using standard forms. The spray operators recorded serial number of the insecticide, date of spraying, where the insecticide was mixed from (compound of the home), and the number of houses sprayed using pre-assigned codes of the insecticide. Additional information captured included household size/occupancy and other malaria control measures used by individuals in each sprayed house.

> “*You just write the form serial number, if the person has agreed for you to spray his or her house, you write yes, ‘Y’ on the form. If the person doesn’t allow you to spray the house, you write ‘N’, No. Then you also write the day when you have sprayed*”--- IDI-20-Spray Operator.

Some individuals in the communities did not allow their houses to be sprayed. However, the heads of these houses were in most cases reported to the local community and district authorities. These houses were eventually sprayed upon follow up by the local authorities. The sprayed houses in the communities were marked with unique codes for ease of identification.

> *You [spray operator] will write everything on that wall and then If the person doesn’t want to write, like this one here, it will remain like that. Then if they [local leaders] come here and then they ask, we’ll just tell them what happened …Even this one the local community leader came here and I told him, this head of this house refused the spray…”* -- IDI-05-Spray Operator.

#### Monitoring quality of mixing of the IRS insecticides and spray exercise

The household heads were encouraged to observe mixing of the insecticides in the communities. Spray team leaders also supervised their teams to ensure that the insecticide was mixed well, and sometimes, the local leaders also joined the spray operators and observed the mixing of the insecticides.

> *“…that medicine in the Sachet (Fludora Fusion) …they first place it in the pump … after they request for some water which they pour in the pump… and they shake after filling then start entering the house and say you have seen me mixing the medicine our names are spoiled here that we spray water”*. --IDI-25-Household Head
>
> *“Make sure as team leader you come there during the mixing of the chemical… you see, is it properly being poured in water? Is it 7.5 liters? If it’s not reaching the 7.5 liters, you say the spray operator should add a little bit of water to come up to 7.5 liters*” --IDI-21-Sprya team Leader.
>
> *“…then you open the sacket you know, you pour in that spray tank. Then after that you close and then you shake it so that it is well mixed, after that you … you pour the rest of the liters of water then after that you close and then you pump the pressure”* -- IDI-19-Spray Operator.

However, some household heads feared to observe the mixing of insecticides because they had no personal protective equipment (PPE) and some did not know what they were observing due to limited information. Some participants suggested community education to help improve monitoring of IRS implementation by the community members.

> “*That was the case, because me going near them for them they were having protective gears, now should I go there minus knowing the instruction? Ah ahh, I have to protect myself by staying away from them, [giving them some distance]*” ---IDI-02-Household Head.

The social behavioral change communication (SBCC) team gave communities information about the benefits of spraying and addressed some complaints therefore increasing willingness to accept indoor residual spraying (IRS). The SBCC team also encouraged the sprayers to allow individuals in communities to observe and monitor mixing of the IRS insecticides.

> *“… and if they are mixing, they will allow the household member or one of the household members to see and know whether it is open or not open, they allow and they give education on how they are going to spray the house…’ ---IDI-15-Team leader or spray operators*.

#### Nature of supervision of the indoor residual spraying

Multi-level supervision of IRS implementation was done; there was supervision at household level as household heads were encouraged to observe the mixing, local authorities, spray supervisors ensured the mixing was done well, spray team leaders who immediately supervised for quality and district authorities including district health assistant who oversaw the entire spray exercise.

> *“We can begin with the team leaders, yes, we had four spray operators being supervised by one team leader. So, the role of the team leader is to make quality. We also had the sub-county, or the site supervisors, they also do supervision. Even at the district, we had s also sub-county task force. All these ones, they were meant to do supervision at that level. At the district we had the district task force, which comprised of the district staff and then the central supervisors…”* --IDI-12-Supervisor.

Physical site monitoring was the main approach to supervision where supervisors and team leaders ensured quality and completeness of spraying. Some supervisors engaged in the spray exercise by supporting spray operators to spray. In the field support visits, supervisors mainly checked for compliance with the standard operating procedures (SOPs).

> *“We moved to various sites on different days to make sure that spray operators are … doing the right work…*”-- IDI-12-Supervisor.

### Opportunities for successful deployment of indoor residual spraying in West Nile region, Uganda

#### Training of spray operators for indoor residual spraying

The IDI participants reported that training received by the spray operators was the main enhancer for successful deployment of IRS. It contributed to quality mixing of the insecticide and ensuring that houses were sprayed well. It also helped the spray teams in overcoming challenges in the field for instance challenges associated with the type of housing.

> “I *think the people who sprayed the house were trained people and they were systematic; they were doing it in an organized way. from ceiling, they now come to the walls, you know mosquitoes like being in hidden places. So, they also targeted the hidden spots where mosquitoes, they believe are hidden…actually the entire house inside part was sprayed*.” --IDI-07-Household Head

Involvement of village health teams (VHTs)/community health workers as spray operators enhanced IRS implementation. This is because the village health teams/community health workers are known within communities and thus it was easy for individuals in households to trust them. The spray operators were given supplies, pumps, polythene to cover household items and bicycles to mobilize and spray households. The spray activity involved multiple stakeholders at different levels including district and community leadership.

> *“Yeah, you know, there are some VHTs who helped us a lot, although they were not all selected to take part in the spraying”* -- IDI-15-Team Leader of Spray Operators

#### Community willingness to have the house receive indoor residual spraying

The respondents indicated that most households were willing to have their houses sprayed which made the spray exercise easier. The community appreciate the fact that they had a mosquito challenge. Some households who had refused the spraying received encouragement from those whose houses were sprayed. Elimination of other house insects was another contributor to the willingness to accept the spray.

> “…people accepted the spray, if anyone *realized they were skipped or missed the followed the spray operators to ask why their house was not sprayed? Therefore most people they accepted this medicine*” --IDI-25-Household Head.
>
> “*Then after there, they came to realize that the chemical is good. Those people who have allowed their house to be sprayed. They will tell you, you see, if you sprayed that, you know my house is now sprayed. This like mosquitoes, houseflies on my house, they are not there. When they heard this the people who had refused the spray later all agreed to have their houses sprayed…”* --IDI-20-Spray Operator

The Social behavioral change communication team helped in management of complaints from communities regarding IRS which further helped in enabling acceptance of the spray for malaria control.

> *“…remember in December 2022 during the first spray using Sachet chemical, there was a rumor that aah three people that had already died from the spray. But we have not started, so that’s how our SBCC team, they went and they managed the whole communities. So, there were those beliefs, but I think in totality they were not so much of a threat. At least our social behavior change communication (SBCC) team, they really convinced and people complied and accepted the spray…” IDI-12-Supervisor*

#### Compliance to the instructions by households during roll-out of indoor residual spraying

During the spray activity in communities, the IRS teams provided guidance to individuals in households. The study respondents indicated that most individuals in households complied with the instructions given by the spray team including, keeping children away from the house during spraying, removing their house items, and keeping outside the house during the spraying.

> “…*what we want to request from people is that you remove all your properties and take outside the house like bed, clothes, food staffs so the chemical doesn’t fall aah like drop on them. …let all the items be collected outside and covered well with polythene bag…then after spraying the medicine they say no one should enter the house they give some time duration I don’t know how long, …”* --IDI-25-Household Head.

The spray teams communicated to household members before and after spraying was done. This helped create good rapport with household members especially through greeting, self-introduction, explaining risk of malaria and the benefits of spraying.

> *“…before you start to spray the rooms for the people. The first thing you must do is, the moment you reach the home of that person, you must get the head of that home. Then after welcoming you, you introduce yourself to that person then you also tell that person why you have come here and what you are going to do. Now after explaining that, you tell the person the importance of the exercise, which is going to take place concerning the medicine…”* -- IDI-13-Spray Operator.

The spray team engaged different stakeholders including district leaders, community leaders including politicians, and religious leaders. This stakeholder engagement at district and community level helped enhance acceptability of IRS.

> *“We also engaged other community members like the parish chief, like the area counsellors, like opinion leaders, like religious leaders. We engaged them. They were willing to support us”* -- IDI-16-Supervisor.

### Challenges faced during implementation of indoor residual spraying for malaria control

#### Hesitancy to accept indoor residual spraying by some households

Some of the respondents reported presence of hesitancy and refusal of IRS among individuals in the community. The study participants reported some of the reasons for the hesitancy including myths about the spray, and misconceptions about the insecticides in addition to cultural beliefs and bias towards government programs. For example, some individuals in the communities had a myth that the insecticide was going to kill people, that it would cause infertility among men, and caused cancer. The communities also had a belief that anew baby should not be brought out of the house for some time like two months which affected the spraying of such houses.

> “*They [people in the community] told us they are going to die because of this Sachet medicine [Fludora Fusion]. So, after that, they told us when somebody refused, we should take him or her to the local authorities (LC1) then the LC1 should refer that person to the sub-county level for the security officer to take action. Some were taken also up to the police afterwards when they returned they called us back to spray their houses…”* ---IDI-09-Spray Operator.
>
> *“The other myth I observed was the women who had delivered, they refused their houses to be sprayed. That they believed that a newborn is supposed to only be moved out of the house after two months. Once the child is taken inside, the child is taken to the bedroom, this child remains there for two months. So, we also faced the challenge where there is a newborn, they refused to move the baby outside…”* --- IDI-16-Supervisor.
>
> *“We had the myths from the communities. Ah people would say that ah because of this insecticide, they cannot be able to produce children*” -- IDI-22-Supervisor.

Some participants said community people were hesitant because they thought governement had wrong motives of reducing the number of their children. Hesitancy and refusal of the spray challenge was addressed through social behavior change communication (SBCC). This helped to make more individuals to accept IRS application in their houses. However, existence of hesitancy and myths in the community is a potential indicator of inadequacy of IRS awareness among communities.

> *“…they [ community people] were saying this chemical has some problems… that it is government plan to kill people. So, I don’t want to spray my house. But we just report to the supervisor, the health assistant and the local security…” -*-- IDI-03-Team Leader of Spray Operators.

Participants in the communities reported instances where individuals smeared the walls of their houses with clay and/or cow dung after the spray. In most homes this was done to remove the strong smell of the insecticide while others smeared their houses as a custom in preparation for celebrations of religious days like Christmas and Idd. It was also noted that some individuals covered walls of their houses with clothes/curtains after the spray to hide the stains left behind by the insecticide.

> *“During Christmas or Eid, they [community people] will say, ah I need to smear my house for it to look very nice, for it to look beautiful. … if these holidays come in between the months when the houses are being sprayed here, the people smear the walls after” ---*IDI-13-Spray Operator.
>
> *“…women like smearing houses …they were told that they were not supposed to smear the walls after the spray. Now, after spraying you find that some also like my neighbor’s, they after spraying, they bring these curtains, they hang them covering, the wall which was sprayed”* --IDI-13-Spray Operator.

#### Inaccessibility of houses for spraying due to absence of household members

Study participants reported that some of the houses were not accessible for the spray as adult members were not present in their homes at the time when the spray team reached. Adult individuals were reported to had gone to work or had social responsibilities away from home. The spray team found only children in some homes which made it difficult to spray such houses as children could make decisions to allow the house to be sprayed.

> *“… no, but the challenge was that the day when they came, some of my tenants were not around. So that also forced the sprayers to come the another day..”-*--IDI-02-Household Head.

#### Difficulty in removing household items and transport of spray materials in the field

In-depth interview participants reported challenges in moving items in and out of the house during IRS deployment. Participants also indicated instances were spray operators supported some households to take out house items in cases of vulnerable occupants like the elderly and the sick. These caused delays in coverage of houses in communities with the spray as it consumed a lot of time.

> *“The first challenge was off-loading all those items outside and then at the same time taking them back inside the house… This really took a lot of time, because I was also having another commitment somewhere…*” ---IDI-02-Household Head.

In the field, participants reported difficulty in carrying spray materials between different houses in the community. This was worsened by not allowing the spray operators to eat while working which made moving from home to home challenging.

> “*You will find that your body drains moving chemicals and tanks for spraying. The sprayers become weak because here you move long distances and also you have to pump the spray. The whole day you will pump the thing, that thing will make you to be weak minus eating”* --IDI-20-Spray Operator.

#### Lack of transport to reach houses in distant areas of the district

The participants reported moving long distances without proper means of transport. Some spray operators had no bicycles and, in some instances, bicycles would not reach some areas like the swampy areas or hills. Some spray operators said they hired bicycles and had to spend some money paying the owners.

> *“Sometimes it’s difficult to carry it[spray pump]. That time they told us to move with bicycle, but I don’t have bicycle at that time…*” -- IDI-06-Team Leader of Spray Operators.
>
> *“We were using bicycles. You know, when going to the field, you find difficulties in riding this bicycle you find these tyres also get spoiled. Also the sunshine was also a lot.. we struggled, and we finished”* -- IDI-03-Team Leader of Spray Operators.
>
> “*Because we, that time we use bicycles yeah. Before we went to the field, we were trained, after training they told us, if you don’t have your bicycle, you are not going to participate in the exercise. So, we all went to look for bicycles. For those who do not have bicycles, they borrowed”* --IDI-10-Spray Operator.

#### Inadequate supplies to support indoor residual spraying

Participants reported inadequate stock of supplies such as spray chemical, polythene to cover household items, and gloves. There was inadequate personal protective equipment for the spray exercise.

> “*Like gloves, we suffered because the gloves were not enough. Like we as team leaders, we were not provided, we just give them to spray operators because we were not close to the drug*” --IDI-08-Team Leader of Spray Operators.

Shortage of the insecticides were experienced in the first spray cycle in November/December 2022, and the second cycle December 2023 in West Nile region. During the spray exercise there were reported instances of theft of the insecticide. Study respondents shared instances where operators were reported to have hidden the IRS insecticide. The chemicals that were hidden were reported to have been used spraying plants like ‘Mairungi Trees’ (Cannabis plant).

> *“There was an experience that we had from these spray operators, some of the communities told us that these people who are spraying houses are hiding some of these drugs. You know, majority of our people here have this tree which we call “marungi”. Somebody has a I don’t know how the test has been done, I heard this thing also worked very well in the spraying of those trees.”* --IDI-02-Community Leader

### Administrative challenges experienced during implementation of indoor residual spraying for malaria control

Inadequate funds for social mobilization and buying of personal protective equipment (PPEs) for the spray operators was reported. There was delay in paying the spray team members including supervisors which became a de-motivating factor.

> *“…Ministry of Health maybe to come up with figures, the entire budgeting was done from that side. So that’s why some areas lacked budget. Besides that, when the budget was declared to us, again the ministry followed up to take some of the money away, which was meant for social mobilization. So, in a way, we had a limited budget to cover the entire district at once”* — IDI-23-Supervisor.

Participants shared payment of their allowance as a challenge as they were not aware of the payment terms and frequency. This was coupled with delays due to bureaucratic systems and some felt that the pay given was little.

> *“…after the exercise, we did not face much challenge except payment. There was delay in payment. So, they took us long to pay our rewards …Like two months”-*-- IDI-16-Supervisor.
>
> *“The payment is not good for us because if you see this work is not easy. At the moment when we start working like this, they do not tell us the money which we are going to be paid…. Then when we are finishing work, that’s the time when they came and give us the forms to fill for money…*” —IDI-09-Spray Operator.
>
> *“I used like 30,000 Uganda shillings (about 9 USD) during the exercise, calling a lot of people even calling direct to site supervisor, calling technicians, calling spray operators, where are you, what is missing, what is working well. So, with all this, I may find my money I get from payment reduced. My bicycle also got spoiled, no repair. I got stucked. Even the money which we are working for delays to come…” --*IDI-21-Team Leader of Spray Operators.

Field maintenance of the spray equipment and other appliances in cases where there is damage was a challenge to the spray teams. Whenever the spray equipment broke down there was no means for repairing them which affected IRS deployment.

> *“And also, the spray tanks sometimes they fail to work. That one also makes releasing the medicine to go on the wall difficult, sometimes the droplets [ehh] come back to you…”* — IDI-13-Spray Operator.

Delayed communication and community sensitization about IRS deployment resulted in resistance to the spray exercise due to rumors.

> *“…it was poor communication other people say it is delayed communication…this communication reaches us late… So, I expected as we used to do in the health centers before we call the local leaders, plus the elder people… tell them about this exercise that we are intending to come and carry in their communities. So, these are the first mobilizers, they will come and mobilize people, tell them on what is coming, so that people are aware…”-*-- IDI-18-Community Leader.

The study participants reported that the number of spray operators were few and this caused inconveniences to people who had to take house items outside as they waited for sprayers who took long to reach them and sometimes, they would not come and postpone to another day.

> *“And the other one was the number of the sprayers, according to me, were not enough. Why I’m saying so is that you could make the LCs to go and mobilize, that you people remove your things out, these people are coming. You remove your things, at times you wait for these people, they could not even come because maybe their number was very few, even others removed their things from their house two to three times. So that way it was not good to me”* ---IDI-02-Household Head.

Leaving out some of the village health team members (community health workers) led to de-campaigning of the IRS program in the communities.

> *“You know, there’s always some allowance attached to the program. When a VHT, like let me now take the experience of this, the current one of the bottle chemical (Actellic). The previous one of sachet (Fludora Fusion) got me when I was in Lodonga. These people here of Kuru sub-County, because we have 35 villages, each village is having two VHT. When you calculate this number, it is almost 70 something. And when the program is coming, it is targeting about 40 spraying operators, leaving out the rest of the VHTs. Because everybody is expecting that small facilitation …If you are left out, it will bring a lot of problem…Instead, some of them now de-campaign the program”* — IDI-04-Community Leader.

Lack of experience in the deployment of IRS was identified as a key challenge during the first spray cycle in 2022 (Fludora Fusion). The participants reported that the IRS exercise of 2022 was difficult because it was the first of its kind in West Nile region hence there were no lessons to benchmark from.

> *“I think aah was really a hard one to begin with, being the first time, I mean being the first IRS, it was not easy. However, we struggled, we put all our energy, we had to do it. I think we even did it successfully*” ---IDI-12-Supervisor.

#### Challenge of the nature of houses in the communities

The sprayers reported that smooth walls did not absorb the chemical as it slide off after spraying.

> *“…because of that softness of the wall, while spraying the insecticide, you could see some medicines drop out or drop down and being poured out because there’s no rough surface to just absorb that medicine there.”* --IDI-21-Team Leader of Spray Operators.

The IDI participants reported presence of sacred artistic drawings on the inside of walls of some houses that people never wanted to get damaged by the spray which made them refuse the IRS.

> *“…There are some houses which had these drawings on the wall… you should not spray the house as it washes away the pictures…*” --IDI-09-Spray Operator

In most rural communities, houses are small in size and built using indigenous traditional techniques and materials like clay. The small size of the houses in communities made it difficult to operate inside to spray the insecticide.

> *“…there are houses which have different structures…Like you might get into a house where the house is too small for one to enter inside and spray … you can spray that house within like even less than 30 minutes…”* -- IDI-03-Team Leader of Spray Operators

### Perceptions regarding the insecticides used for indoor residual spraying in the communities

The study participants reported that it was easy to confirm that Actellic (bottle) was sprayed because it left behind an odor unlike Fludora Fusion (Sachet). Some participants indicated that that water could be sprayed during Fludora Fusion IRS, and you wouldn’t notice.

> *“I would prefer Actellic…when you consider Fludora…. If somebody sprays water, like… what some community members were alleging, if somebody sprays water, you cannot tell if Fludora was sprayed*” ---IDI-12-Supervisor.

The participants stated that it was easy to mix Actellic as compared to Fludora Fusion as Actellic was in liquid form in a bottle which you just pour into the tank without need to shake to dissolve.

> *“I was saying they should continue to use this recent one (Actellic) because that one is just in the bottle. You can just pour it in the tank and it dissolves. But Fludora, you have to shake vigorously before spraying…”* --IDI-10-Spray Operator.

However, the strong smell and the fact that Actellic supply was not adequate were the main limitations that faced spraying of Actellic.

> “*But the one for 2023 (Actellic), outside the house there are no mosquitoes. The problem with it is the smell*”---IDI-06-Team Leader of Spray Operators.
>
> “*Like at first, 2022 (Fludora Fusion), we sprayed the houses successfully. Everybody got the chemical but last year, 2023 (Actellic) the chemical was not enough. Some parishes missed the chemical*” ---IDI-08-Team Leader of Spray Operators.
>
> *“Yeah, of 2023 (Actellic), the number of the chemical was not enough…even now as I’m talking, the other houses in my cell they are not sprayed…they have missed it… not here in the town council alone the whole of the district even you go to other sub counties. The whole village is not to sprayed…. because the number of medicines they are few”*-- DI-24-Supervisor.

The participants reported that Actellic was perceived to be more long lasting compared to Fludora Fusion by communities.

> *“This one of Actellic for me, it is good.… Even right now, you can’t hear that voice of mosquitoes, if you open the door like this even you still smell that chemical. The stain is still there on the wall because that one stays for nine months on the wall. This one, this Actellic stays for nine months but the other Sachet one (fludora) stays for like six months”* --- IDI-19-Spray Operator.

## Discussion

In communities, indoor residual spraying (IRS) was observed to have an effect in eliminating mosquitoes in sprayed houses for a given time depending on the insecticide which was used. This in addition to killing other household insects like cockroaches, houseflies and ants and appreciation of the existence of the malaria problem in the communities motivated individuals to accept the spray. This is like findings of previous studies in Mozambique (20), Tanzania (21) and Uganda (22, 23). Perception of effectiveness remains a key driver of IRS acceptance for malaria control (22, 24). This gives malaria control programs in the country an opportunity to scale up IRS in the fight against malaria. The study participants also reported involvement of the social behavior change communication (SBCC) team in handling complaints against IRS in communities. This in addition to the inclusion of local community members like the village health teams in the spray exercise further enhanced acceptance. The finding of the key role played by SBCC and inclusion of known members of the community in acceptance of IRS has implications for future spray implementation. Local, and transparent recruitment of spray teams thus remains a key consideration in improving IRS acceptance for malaria control (24).

Although community wide IRS education was not mentioned by the IDI participants, training of the spray teams prior to spray deployment was reported by most respondents in this study. The spray teams were aware of the need to protect individuals from getting exposed to the IRS insecticides. This was indicated by the instructions the spray operators provided to households prior to the spray including keeping children away, closing all doors and windows during and after, not entering the house immediately, covering household items, and avoiding contact with the sprayed walls. However, challenges including shortage of supplies like personal protective equipment (PPE), and polythene bags for covering household items persisted. An indicator of the complex logistics in the implementation of IRS (4). Previous studies have indicated the significance of community education on IRS uptake (22, 23) and is also recommended by the World Health Organization (15). Strengthening of community education thus remains a key consideration in subsequent IRS implementation for malaria control in the country. However, these activities need to be conducted ahead of time to give communities time to adjust. In the case of West Nile community engagement and training were not done in time and resulted in some houses missing the spray as the owners were not around since the exercise found them when they were not around. This could compromise the effectiveness of indoor residual spraying in malaria mosquito control.

In this study cultural beliefs and myths regarding IRS such as causing infertility and death among exposed individuals were reported in communities. Study respondents indicated that houses were there was a newborn baby declined to allow for spraying as culture did not allow the newborn baby to be brought out of the house for some time. In addition, participants reported hesitancy towards IRS due to the fear of affecting their reproduction or fertility especially causing impotence among men. This is like findings of a previous study that reported community hesitancy towards IRS driven by fear of adverse effects of the insecticides, cultural beliefs, and attitudes (21, 22). Additionally, a previous study in Mozambique found that indoor residual spraying was the least preferred malaria control measure during pregnancy (25). Hesitancy among households coupled with operational challenges could potentially hinder community IRS coverage and the effectiveness in malaria control.

In the communities, study participants commonly reported practices like smearing surfaces of inside walls of sprayed houses with clay or cow dung, and covering of sprayed walls using clothes/curtains. In addition, absence of adult individuals in homes during time of the spray and difficulty in removing house items were mentioned by study respondents. The challenge of removing house items potentially impacts community coverage of IRS and has been reported by previous studies as a key barrier to IRS implementation (18, 22). In our study this was further exacerbated by postponement of the spray exercise in some cases after individuals had removed their house items. Individuals smeared or placed curtains on sprayed walls reportedly to reduce the smell and stain on the walls left behind by the insecticide. Also, smearing walls of houses using clay or cow dung was reported in preparation for festive sessions or religious holidays a common practice in Uganda especially in rural settings. Use of IRS insecticides that left behind a strong smell or stain on the walls have been reported in previous studies as a barrier to spray implementation(18). These finding of local practices in households in rural settings has implications on community engagement and education in subsequent IRS implementation.

Some participants reported instances where individuals did not allow spray operators to apply IRS in their houses, however, the intervention of local authorities helped ensure that these houses receive the spray. This was enabled by the compliance of communities to the local authorities a finding like that of a previous study in Ghana by Suuron et al., (26). The spray operators were in some cases also hindered from entering houses in rural settings due to the small size of the house. The IDI participants in this study reported difficulty in operating inside such structures. With malaria being most prevalent in rural poor settings in most sub-Saharan Africa countries, the nature of houses in these communities is thus a potential barrier to the deployment of IRS in malaria control.

In Uganda one cycle of IRS is deployed annually where Yumbe district has currently received up to five cycles since 2022. Annual implementation of IRS for malaria control is adopted by most malaria affected settings globally irrespective of the residual efficacy of the insecticides (8). In this study participants perceived the insecticide in the sachet (Fludora Fusion) to have residual efficacy of about six months, and the bottle (Actellic) was said to be effective for up to about nine months. While this may not reflect the actual residual efficacy of the insecticides used, previous studies have reported 5-6 months residual efficacy of Actellic and Fludora Fusion (7). The annual deployment of IRS irrespective of the residual efficacy could thus impact its effectiveness for malaria control. This is further exacerbated by the rise in mosquito vector resistance (13) and the change in mosquito composition and behavior following sustained implementation of IRS (7, 27). The study participants reported challenges in mixing of insecticides, this coupled with reported inadequate IRS supplies is likely to further compromise the quality of the spray and thus the annual deployment.

The study had some limitations as data was collected from individuals of the community who participated in the two rounds of indoor residual spraying in West Nile region, it was not possible to confirm how individuals recruited into the study were involved in the IRS activity. This was especially challenging as there was no documentation of their engagement. However, this study included stakeholders at different levels including district, sub-county, parish and village level. This helped in triangulating the experiences of the study participants and improve the validity of our findings.

## Conclusion

Communities were highly receptive towards IRS for malaria control with most study participants being positive on its effectiveness. Training of spray operators, involvement of different stakeholders, community compliance with local authorities, and involvement of social behavior change communication (SBCC) help enhance implementation of IRS. However, the deployment of the spray faced some challenges including inadequate supplies, removing and returning household items, and difficulty moving spray materials in the field. The Ministry of health should prioritize strengthening timely community engagement prior to IRS implementation.

## Data Availability

All the data for the study are provided within the manuscript.

## Acknowledgments

The efforts of Olwortho Wilfred and Obeida Tabuga the social scientists who conducted field interviews for collection of data for the study. We also acknowledge the study participants and district leaders who help in enabling the study team access individual participants in communities for the in-depth interviews.

